# Passive sensing data predicts stress in university students: A supervised machine learning method for digital phenotyping

**DOI:** 10.1101/2023.07.29.23293375

**Authors:** Artur Shvetcov, Joost Funke Kupper, Wu-Yi Zheng, Aimy Slade, Jin Han, Alexis Whitton, Michael Spoelma, Leonard Hoon, Kon Mouzakis, Rajesh Vasa, Sunil Gupta, Svetha Venkatesh, Jill Newby, Helen Christensen

## Abstract

University students are particularly susceptible to developing high levels of stress, which occur when environmental demands outweigh an individual’s ability to cope. The growing advent of mental health smartphone apps has led to a surge in use by university students seeking ways to help them cope with stress. Use of these apps has afforded researchers the unique ability to collect extensive amounts of passive sensing data including GPS and step detection. Despite this, little is known about the relationship between passive sensing data and stress. Further, there are no established methodologies or tools to predict stress from passive sensing data in this group. In this study, we establish a clear machine learning-based methodological pipeline for processing passive sensing data and extracting features that may be relevant in the context of mental health. We then use this methodology to determine the relationship between passive sensing data and stress in university students. In doing so, we offer the first proof-of-principle data for the utility of our methodological pipeline and highlight that passive sensing data can indeed digitally phenotype stress in university students.

## INTRODUCTION

Stress is the physiological and psychological response when environmental demands outweigh an individual’s ability to cope (Cohen et al., 2007). Although brief exposures to low levels of stress are normal and even possibly beneficial for performance (Yuen et al., 2009), chronic stress can lead to serious consequences including depression, burnout, and disease (Bianchi et al., 2014; Cohen et al., 2007; Melamed et al., 1992; Weber & Jackel-Reinhard, 2000). Early adulthood is characterized by rapid changes, both physiological and psychological. Further, there are many new challenges during this time including changing environments (e.g. moving to a new city), deadline pressures, increased social interaction and balancing school and work demands. Combined, these changes and pressures make university students particularly vulnerable to stress (Hamaideh, 2011; Lu, 1994; Verger et al., 2008). They are well-documented to develop signs of stress-induced psychological distress, which can lead to low academic achievement, interpersonal problems, depression, burnout, self-harm, and suicide (Ribeiro et al., 2018; Sharp & Theiler, 2018).

Numerous mental health self-help mobile applications have been developed to combat university students’ poor mental health (Neary & Schueller, 2018). Here, students who are feeling distressed can rapidly access affordable, or free, applications that aim to provide them with coping skills and strategies for self-help without the need for traditional face-to-face psychological sessions with a therapist. Further, mobile phones are increasingly accessible and popular for younger people and therefore present an easy tool by which to deliver digital health services (Holtz et al., 2020). In line with this, younger adults show substantial interest in trying smartphone mental health apps, a phenomenon that was further increased by the recent COVID-19 pandemic (Ahuvia et al., 2022; Bautista & Schueller, 2023; Montagni et al., 2018). Students, in particular, are attracted to these types of apps due to immediate availability, convenience, confidentiality, and an ability to avoid the stigma associated with seeking face-to-face appointments with mental health specialists (Holtz et al., 2020; Kern et al., 2018). Additionally, students experiencing academic stress and burdens associated with transitioning to post-secondary institutions are known to seek out smartphone apps to help them cope (Melcher et al., 2022).

Mental health smartphone apps have the benefit of being able to collect copious amounts of data from single users. Importantly, smartphones have multiple passive sensors that enable tracking of various aspects of users’ day to day lives. This passive sensing data includes GPS, which determines the location of the phone, an accelerometer and gyroscope to measure the acceleration in space and patterns of physical movement, and a step detector to estimate the number of steps. There has been a growing interest in using this passive sensing data to identify variables that can predict mental health status and outcomes. Variables that have been extracted from passive sensing data have been shown to be associated with mental health and psychiatric disorders including depression (Saeb et al., 2015), stress (Ben-Zeev et al., 2015), dementia (Galambos et al., 2013), bipolar disease (Beiwinkel et al., 2016) and schizophrenia (Ben-Zeev et al., 2016).

Despite the growing interest in the relationship between passive sensing data and mental health, little is known about its relationship with stress in university students. Stress in and of itself is an important adaptive mechanism of survival that helps the body to mobilize resources to respond to threat. However, the chronic activation of the stress response system can lead to catastrophic physical and mental health outcomes. More specifically, chronic stress has been shown to lead to depression (Sawatzky et al., 2012), and problems with cardiovascular (Steptoe & Kivimäki, 2013) and immune (Winsa et al., 1991) systems. Smartphone-based interventions and tools that efficiently diagnose stress early are urgently needed to prevent these substantial health burdens.

Additionally, although previous research has shown a relationship between passive sensing data and mental health outcomes, there has been limited description of the methodologies used to both extract and process this type of data. For example, GPS data often has differences in the accuracy of determining coordinates due to a high dependency on factors including unobstructed receivers and good reception. Another example is that irregular smartphone internet connections can result in a data loss. It is imperative, therefore, to begin to work toward establishing clear methodologies that can translate into reproducible research using passive sensing data.

In this study, we had two central aims: (1) to establish a clear methodological pipeline for processing passive sensing data and extracting features that may be relevant in the context of mental health and (2) to use this methodology to determine the relationship between patterns of university students’ mobility, as indicated by passive sensing data, and their stress levels. In doing so, we offer the first proof-of-principle data for our methodological pipeline and, using supervised machine learning models, demonstrate that passive sensing data can indeed digitally phenotype stress in university students.

## METHOD

### Study Design and Participants

Mental health app user data was collected from the Vibe Up study (Huckvale et al., 2023). Vibe Up is a data collection application built for Android and iOS that uses an artificial intelligence algorithm to deliver the most effective mental health interventions to university student users in Australia. Participants of this study also completed survey-based mental health and wellbeing assessments throughout. Passive data, including accelerometer, gyroscope, activity monitoring, distance, and step count was collected across all 30 days of the study. Eligibility for participation was defined by the following: ≥ 18 years of age, currently attending a tertiary institution in Australia, remaining in Australia throughout the study period, and completed screening surveys. Users also had to have a Kessler Psychological Distress Scale (K10) score of <20 (Andrews & Slade, 2007) and Suicidal Ideation Attributes Scale (SIDAS) (van Spijker et al., 2014) >21, to ensure that although users weren’t likely to be “well” they didn’t have a high level of suicidal ideation. The study was approved by the University of New South Wales Human Research Ethics Committee, approval no. HC200466.

### Questionnaires

At screening, users were asked questions about their demographic information including age, sex at birth, sexual orientation, language spoken at home, international or domestic student status, previous mental health diagnosis, and whether they used online mental health services in the past 12 weeks. Once users started the Vibe Up app, they completed the Depression and Stress Scale (DASS) three times across the study (Lovibond & Lovibond, 1995). Here, responses are encoded using 4-item rating scale ranging from ‘Did not apply to me at all’ (0) to ‘Applied to me very much, or most of the time’ (3). As stress was our primary outcome of interest (or output variable), we only used responses to the stress subscale. The level of stress was determined by summing the item scores, multiplying it by two, and converting it to a z-score using reference values for the mean (11.19) and standard deviation (8.25) of the general population of young adults aged 20-29. Further, participants were discretised based on the z-score into no stress group (z-score < 0.5), mild to moderate (z-score 0.5 to 2.0) and severe to extremely severe (z-score > 2.0) as has been done previously (Parkitny & McAuley, 2010).

### Passive Sensing Data Collection

Passive data collection is managed by the Conductor Software Development Kit (SDK), which collects data based on predefined schedules. For Vibe Up, this collection period was all day for 30 days (the duration of each trial). The sample rates of each stream are unique based on the data being collected. GPS location is only recorded whenever a user has significantly moved. Similarly, activity monitoring, distance, and step count are only recorded if a user is actively moving. Accelerometer data is continuously recorded at 50 Hz on iOS and 60 Hz on Android. Gyroscope is recorded at 50 Hz, but on iOS this can only be collected while the app is in the foreground.

### Passive Sensing Data Feature Selection

The use of passive sensing data is becoming more widespread in the literature on mobile app use. Despite this, there remains little consensus and few, if any, descriptions of the methods by which passive sensing data is processed and features are extracted for downstream statistical and predictive models. Here, we suggest an approach that focuses on building directed, edge-weighted graphs that capture the main features of user mobility patterns (Fig. 1).

**Figure 1.**
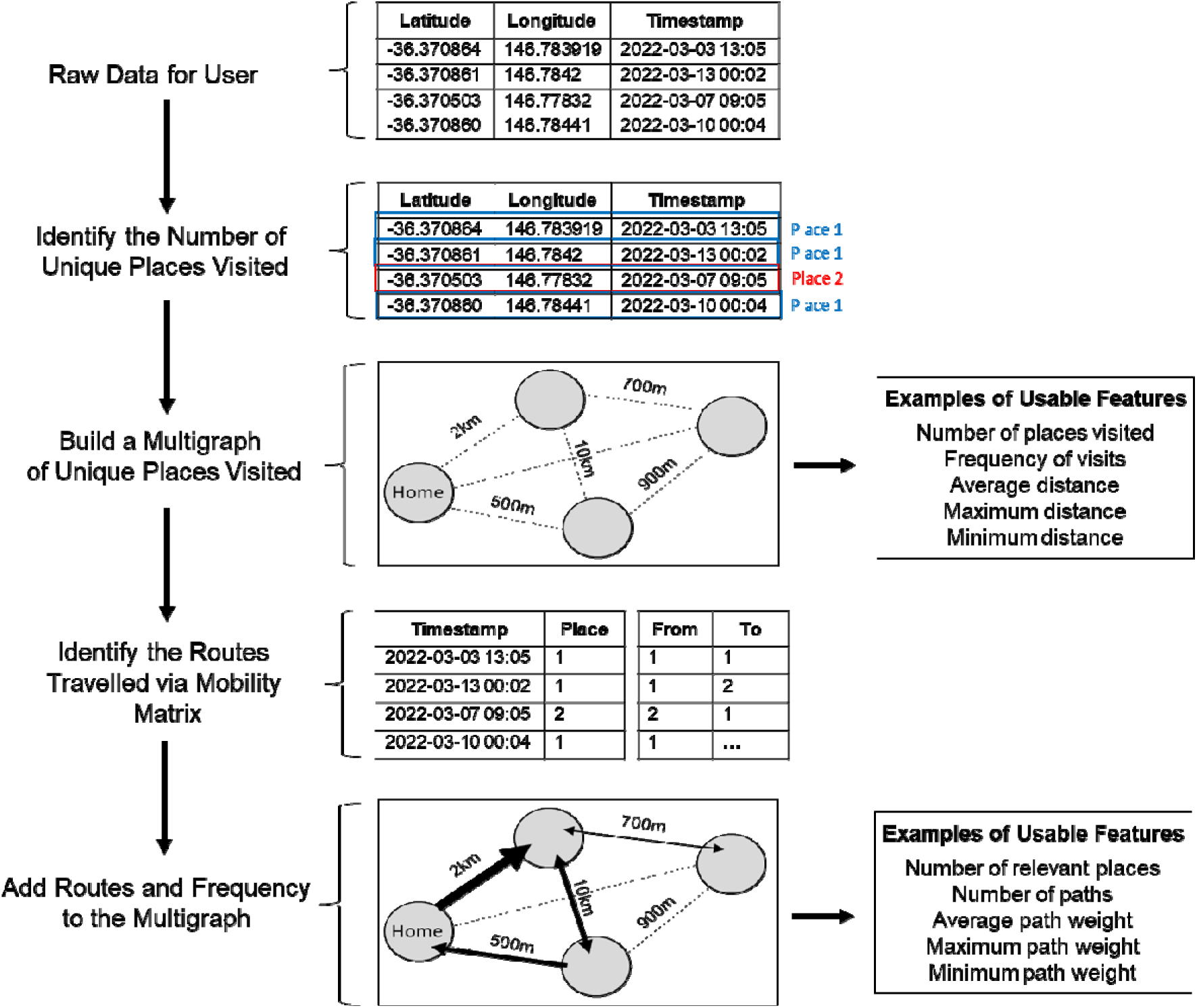
Schematic of the steps used to process raw passive sensing data collected from users’ mobile phones and identify usable features for subsequent statistical analyses and predictive modelling.

#### Stage 1: Processing Raw Passive Sensing Data

The first consideration is that GPS coordinates collected from mobile phone apps can have technical inaccuracies that cause a specific location to look different every time an estimation occurs (i.e. has slightly different GPS coordinates). Therefore, we present each location as a multigraph with area of 5000m^2^, considerably larger than the average property size in Australia, to reduce noise that may be caused by these inaccurate coordinates and randomly captured activities while someone is moving around their property. All coordinates that fall into this area are considered as one location. From this data, it is then possible to: (1) estimate the number of places a user visits, (2) calculate the distance between these places, identify which location is likely to be home (highest number of occurrences / visits), and identify which locations are likely to be irrelevant (lowest number of occurrences / visits). This data processed at this first stage can then be translated into usable features for statistical analyses and/or predictive models, including: number of places visited, frequency of visits, average distance, and maximum and minimum distances.

#### Stage 2: Characterize the Routes and Paths Between Places Using Mobility Matrices

After identifying the number of unique places visited, we can then use the corresponding GPS timestamps to compute a mobility matrix for each user. The mobility matrix, therefore, contains information regarding date of visit, time of day (from 00:00 to 23:59), and the order of visited places at specific time intervals daily across the study. From here, we can then calculate routes and paths that a user has taken between places. Further, by using the number of passes between visited places (used to determine how often the route is used), computed in Stage 1, we can estimate the relevance of the routes between different places as well as the direction of travel. Importantly, unlike in Stage 1, Stage 2 data processing covers most aspects of human mobility: how many places a user visits, how many routes a user uses to arrive at those places and how often, the typical order of places visit, the time the working day starts and ends, average time a person spent at a particular place, what the night time lifestyle looks like (e.g. frequent night activities suggesting socialization), and so forth. Additionally, it is worth noting that some apps collect data about the types of movement, number of steps, and distance travelled. Once overlapped with the GPS dataset, it is possible to add this data to the geometric maps to determine the preferred way of travelling between places. Although we did attempt to collect this type of data in the present study, there was a lack of overlap between steps and GPS timestamps. Therefore, instead of merging these data together, we treated steps as a separate variable and summed up the number of steps per day. We then used quantiles (25%, 50%, and 75%), as well as maximum and minimum values as features. Overall, the data from Stage 2 can be extracted as several features including number of relevant places, number of paths, average path weight, and minimum and maximum path weights.

### Analytical Approach

To determine if the extracted passive sensing data features could predict stress, we deployed several machine learning algorithms including general linear algorithms (lasso and ridge regression, shrinkage discriminant analysis (SDA)), geometric distance-based algorithms (k-nearest neighbour (KNN)), tree-based algorithms (classification and regression trees (CART), random forest), and artificial neural networks (ANN). The dataset was split into a training dataset and testing, held-out dataset (70% and 30% respectively). Machine learning models were built, fine-tuned and validated on the training dataset by using three-fold cross-validation repeated five times. For the final assessment of machine learning models’ performance a held-out dataset was used. Where there were class imbalances of the output variable, an oversampling technique was used whereby the underrepresented class is randomly resampled to ensure that the algorithms receive approximately the same number of classes. For all algorithms, a fine-tuning grid method was used where all possible combinations of parameters within the predetermined ranges were estimated (Table 1).

**Table 1.** Parameters of the supervised machine learning algorithms used to predict stress based on passive sensing data features.

| Model | Parameters |
| --- | --- |
| Linear regression | $\alpha$: 0, 1<br>$\lambda$: 0.001-1 |
| Shrinkage discriminant analysis (SDA) | Diagonal: true, false<br>$\lambda$: 0.001-1 |
| K-nearest neighbour (KNN) | Number of the nearest neighbours: 1-15 |
| Classification and | Complexity parameter: 0.001-0.1 |
| regression trees (CART) |  |
| Random forest | Number of variables randomly sampled: 1-# of features in dataset |
| Artificial neural networks (ANN) | Number of hidden layers: 0 and 1<br>Number of neurons in hidden layers: 3-50<br>Activation function: tanh, relu, leaky relu<br>Optimization algorithm: Adam, RMSprop, SGD<br>Learning rate: 0.01-0.00001<br>Batch size: 16-32<br>Epochs: 10-100<br>Regularization dropout layer with probability: 0.1-0.8<br>Kernel regularizer: L1, L2 |

To estimate the performance of the binary classification models, we used area under the curve (AUC). This measure reflects the level of sensitivity and specificity of the model and thus general distinguishing capacity of the model. We also used precision, indicating the proportion of positive predictions is correct, recall, to indicate the proportion of actual positive cases that were predicted correctly and F1, which is a harmonic mean of precision and recall.

All inferential statistics were performed using Kruskal-Wallis followed by a post-hoc Dunn test for three samples comparisons. A Benjamini-Hochberg multiple correction was applied to adjust the *p* values and reduce the risk of a false positive. To determine the correlation between features and the output variable, a Pearson correlation coefficient was used. Analyses were performed in RStudio with R 3.6.3. Supervised machine learning was done using the caret package and neural networks were built using keras library.

## RESULTS

### Participant Characteristics

The demographic characteristics of the Australian university student users of the Vibe Up app are shown in Table 2. The average age of user was 23.6 (range 18 to 34). The majority of users identified as female (76%), spoke English at home (94%), and were domestic students (95%). Approximately half of the user group had a previous mental health diagnosis (54%) although comparatively fewer (25% of users) had used online mental health services in the past 12 weeks. A substantial number of users (39%) identified as being LGBTQIA+ (Table 2).

**Table 2.** Demographic characteristics of the university student users of the Vibe Up app

|  |  |
| --- | --- |
| Sample numbers | 409 |
| Age (average, ± SEM, and range) | 23.6 ± 5.2 (18-34) |
| Sex at birth | 76% female |
| Identify as LGBTQIA+ | 39% |
| Speak English at home | 94% |
| Domestic student | 95% |
| Previous mental health diagnosis | 54% |
| Used online mental health service in the past 12 weeks | 25% |

### Features show weak relationship with stress z-scores

We first performed inferential statistics determine which features, if any, show significant relationship with the output (Fig. 2).

**Figure 2.**
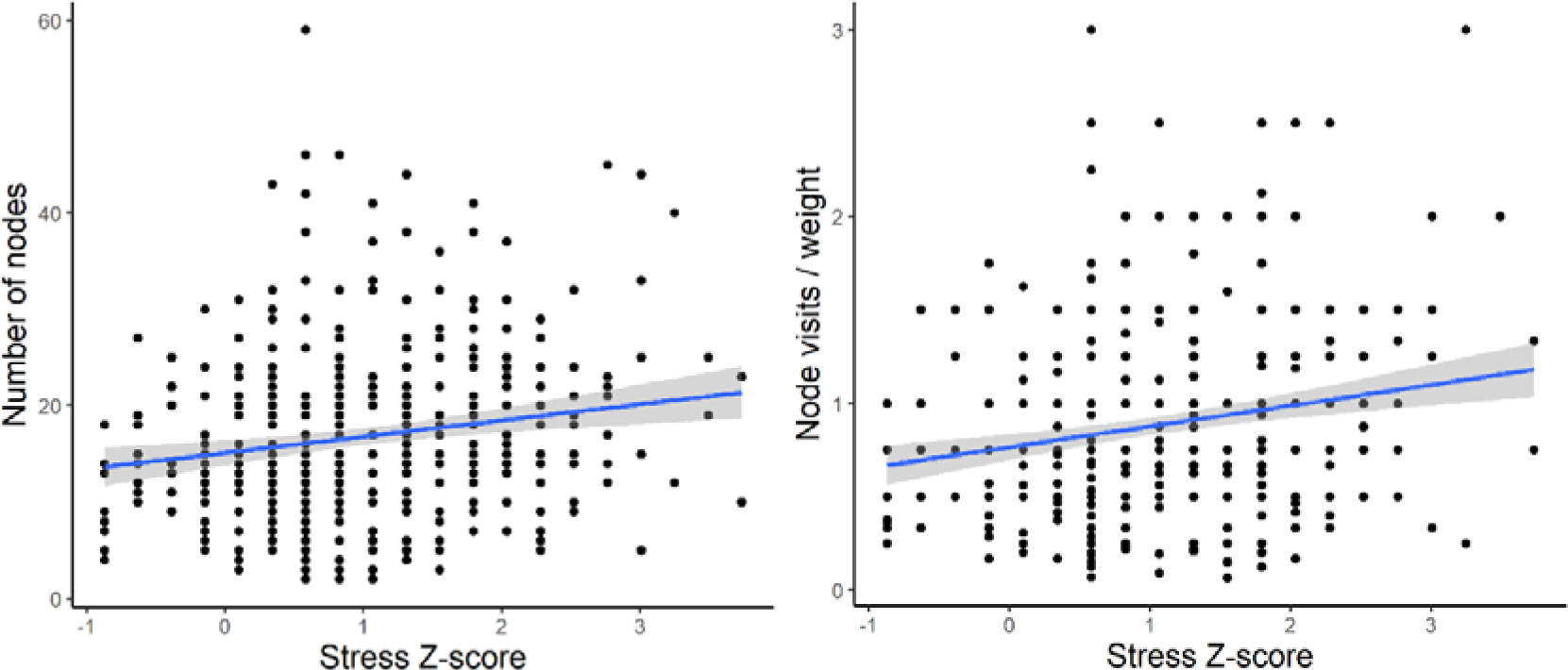
Examples of the linear nature of the relationship between passive sensing features and output (stress) for the best performing features. (A) Individual feature of number of unique nodes (r = 0.17). (B) Engineered feature of combined average node visits per day with 75^th^ quantile of graph weight (r = 0.21).

Although twenty features significantly correlated with stress z-score, the correlation coefficients (-0.15-0.17) indicated a weak relationship between the features and output. To further confirm our initial finding, we used linear regression with number of unique nodes as a predictor and the stress z-score as the value. Similarly, although the overall fit was statistically significant (*p* < 0.0001) it demonstrated a very low R^2^ value (R^2^ = 0.04, F=7.56) and the data points were scattered, indicating that the model is unable to explain the variance in stress z-score. We then tried to perform feature engineering, including weights of graphs, whereby we combined existing features together to examine the correlation with output.

Although this slightly improved the correlations, they were still weak (-0.19-0.21). We further confirmed this by developing and training a ridge linear model on a training dataset and testing the model on a held-out dataset. This resulted in an RMSE of 0.87, suggesting that the model was misclassifying by an entire category of users.

Our initial findings highlighted that weak correlations don’t result in predictive power. One way to improve the performance of our predictive models is to discretise the output variable into a few groups. This would shift away from a regression-type problem towards classification.

### Multi-class classification is similarly unable to predict stress

We binned the stress z-scores into three categories: no stress (<0.5), mild to moderate (0.5 to 2), and severe to extremely severe (>2). To determine if the use of stress as a continuous, rather than categorical, variable was affecting the ability to develop predictive models, we next binned the stress z-scores into three categories: no stress (<0.5), mild to moderate (0.5 to 2), and severe to extremely severe (>2). A Kruskal-Wallis (χ^2^ = 11.03, df = 2, *p* = 0.004) with post-hoc Dunn test confirmed that there was a statistically significant differences in the number of unique nodes between these groups (Fig. 3).

**Figure 3.**
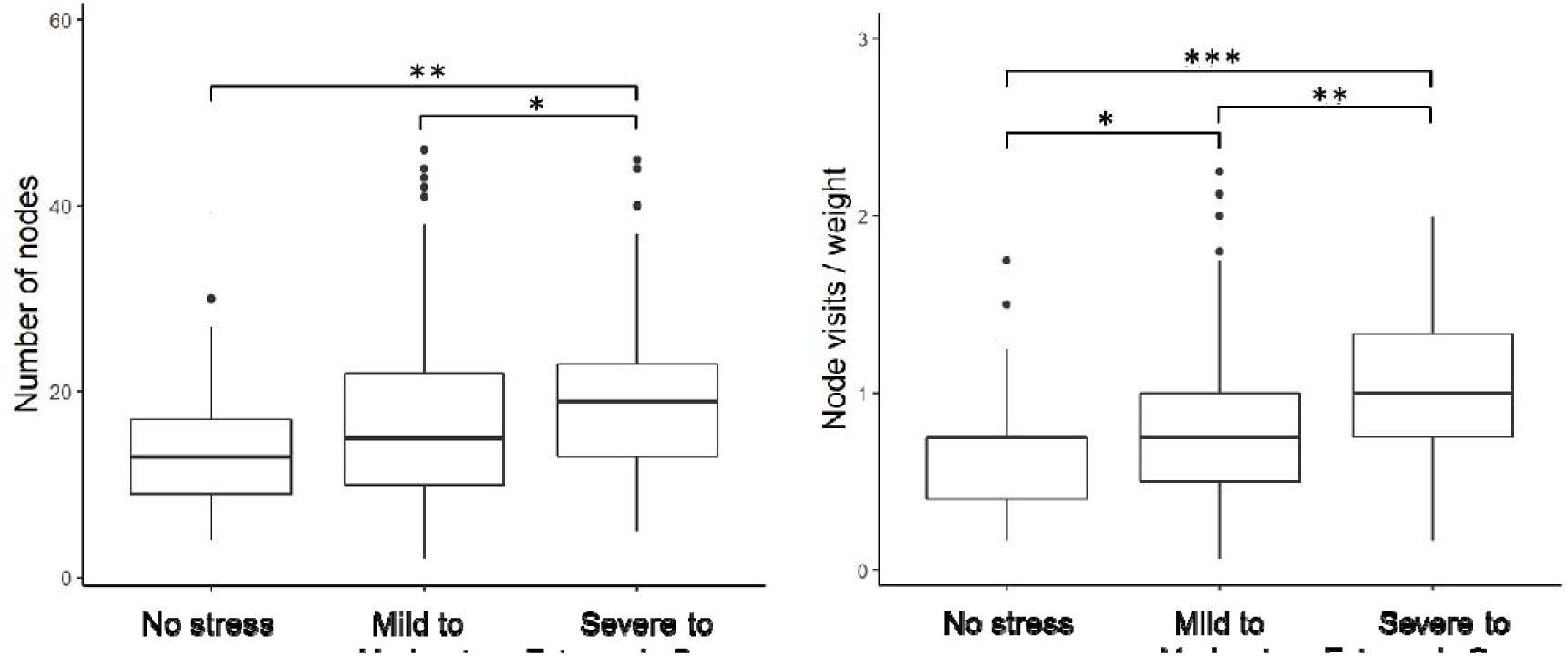
Stress z-scores binned into three categories: no stress (<0.5), mild to moderate (0.5-2), and severe to extremely severe (>2).

After successfully binning the stress z-scores into three distinct groups, we next developed a multi-class classification model, CART, to identify if any of the features were now able to predict stress. The model, however, demonstrated very low predictive power (AUC <0.5).

### The mild-moderate stress group impacts predictive power in a binary classification

We next sought to identify the potential source of our models’ low predictive power. One possibility was the inclusion of the mild to moderate stress group. The rationale for this was twofold. First, there is evidence that mild to moderate stress can be beneficial, including improving performance and efficiency on dual tasks (Beste et al., 2013) and concentration (Degroote et al., 2020). It may be the case, therefore, that while some university students may find stress overwhelming others may benefit from mild to moderate stress. This possibility, therefore, suggests that the mild to moderate group is likely heterogenous and highly variable. Evidence for this can also be seen in Figure 2 whereby the variability in number of nodes and node visits per weight is higher in the mild to moderate users (z-score 0.5 to 2).

There is also substantial overlap between the mild to moderate group with the no stress and severe to extremely severe groups on these two passive sensing measures (Figure 3A,B). An additional consideration was more severe cases of stress in university students are likely to co-occur with clinical mental health diagnoses, including major depression (Muscatell et al., 2009). Therefore, we wanted to assess whether we could improve the clinical translatability by identifying whether digital phenotyping via passive sensing data could differentiate the not stressed from the severely stressed.

After removing the mild to moderate group, we re-performed a binary classification to see if our passive sensing features could predict whether a user had no or severe to extremely severe stress. Using inferential statistics, we first demonstrated that removal of the mild to moderate group improved both the correlation coefficient and significance of the relationship between features and stress (Table 3).

**Table 3.** Examples of features correlated with the stress z score in original dataset and trimmed dataset with no mild to moderate stress group.

| Features | Original Dataset<br>with Three Groups |  | No Mild to<br>Moderate Group |  |
| --- | --- | --- | --- | --- |
|  | Correlation<br>Coefficient | *p* value | Correlation<br>Coefficient | *p* value |
| Number of unique nodes | 0.17 | 0.002 | 0.37 | 0.0001 |
| Number of nodes visited per day (75<sup>th</sup><br>quantile) | 0.14 | 0.003 | 0.36 | 0.0002 |
| Average number of steps per day | -0.14 | 0.003 | -0.25 | 0.002 |

Given that we were able to substantially improve the correlation coefficient, we then deployed several predictive machine learning models to determine if these correlations were sufficiently strong enough to be good classifiers. Using the AUC metric, our models showed satisfactory performance (Table 4). Despite this, however, the precision and/or recall for all but one models was low, indicating that the models struggled to predict at least one of the two groups. We then developed and deployed a neural network to help overcome this and were indeed able to improve the performance metrics, suggesting that it was able to successfully distinguish between users who were not stressed and those who were severely to extremely severely stressed.

**Table 4.** Machine learning algorithm performance metrics for predicting users with no stress vs. severe to extremely severe stress.

| **Machine Learning Algorithm** | **Precision** | **Recall** | **F1** | **AUC** |
| --- | --- | --- | --- | --- |
| Classification and regression trees (CART) | 0.17 | 0.50 | 0.26 | 0.68 |
| Random Forest | 0.24 | 1 | 0.38 | 0.67 |
| Shrinkage discriminant analysis (SDA) | 0.47 | 0.67 | 0.55 | 0.69 |
| General linear model | 0.24 | 0.80 | 0.36 | 0.67 |
| K nearest neighbour | 0.41 | 0.70 | 0.52 | 0.70 |
| Neural network | 0.82 | 0.78 | 0.80 | 0.79 |

### Further polarizing the no stress and severe to extremely severe stress groups continues to improve predictive power of binary classification models

Although it was clear that the mild to moderate group was indeed affecting the predictive power of our models, only our neural network (out of 6 different supervised machine learning models) performed to a sufficiently high level. This suggested that our findings may be limited with respect to generalizability. To address this, we further increased the minimum z-score of the severe to extremely severe stress group (increased z-score of >2 to z-score of >2.2). Again, the rationale for this was that we wanted to determine if there was a potential for clinical translatability of our model to digitally phenotype those who are not stressed relative to those who are experiencing such severe stress that they are high risk of comorbid clinical mental health diagnoses like major depression.

We first confirmed that increasing the minimum z-score of the severe to extremely severe group to 2.2 had a positive effect on the correlation coefficient and statistical significance between our features and groups (Table 5).

**Table 5.**
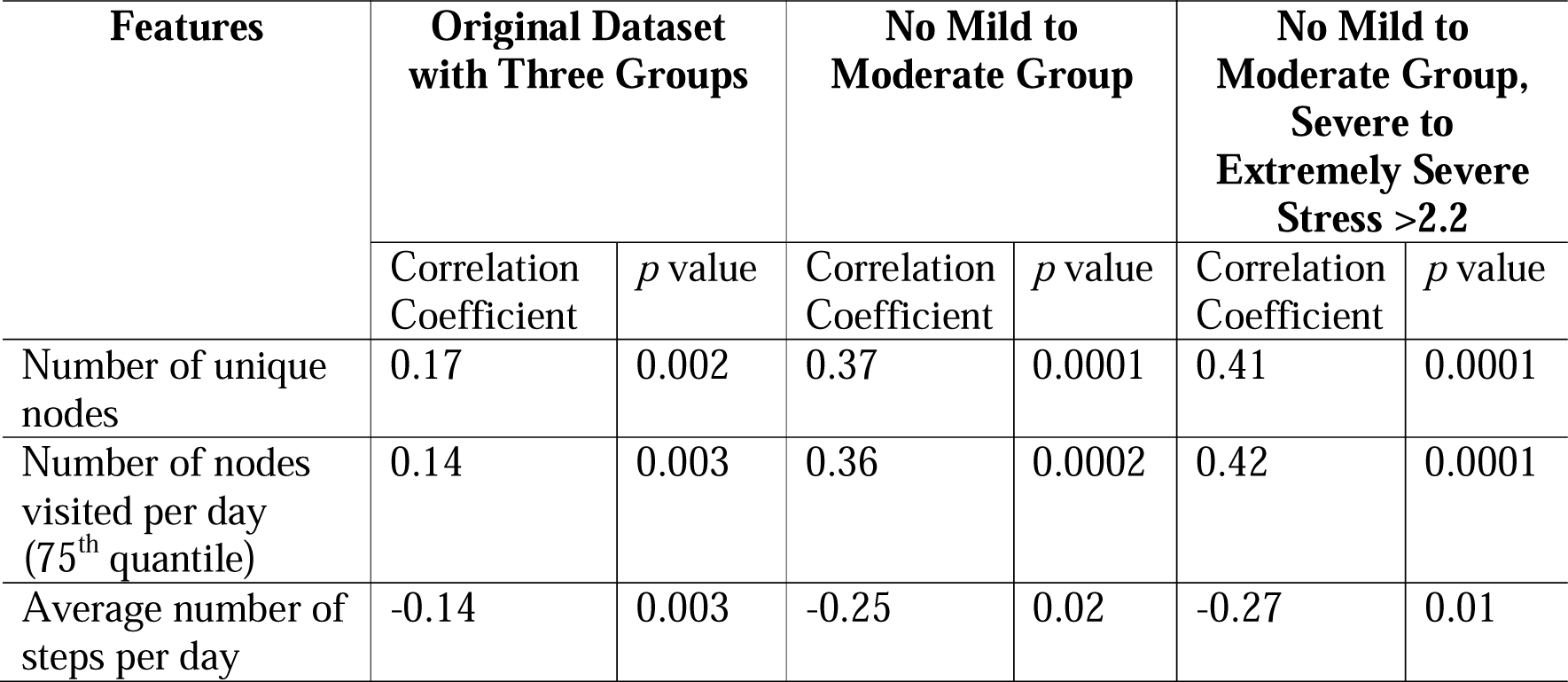
Examples of features correlated with the stress z score in original dataset, trimmed dataset with no mild to moderate stress group, and a dataset where severe to extremely severe stress >2.2

| Features | Original Dataset with Three Groups |  | No Mild to Moderate Group |  | No Mild to Moderate Group, Severe to Extremely Severe Stress >2.2 |  |
| --- | --- | --- | --- | --- | --- | --- |
|  | Correlation Coefficient | *p* value | Correlation Coefficient | *p* value | Correlation Coefficient | *p* value |
| Number of unique nodes | 0.17 | 0.002 | 0.37 | 0.0001 | 0.41 | 0.0001 |
| Number of nodes visited per day (75<sup>th</sup> quantile) | 0.14 | 0.003 | 0.36 | 0.0002 | 0.42 | 0.0001 |
| Average number of steps per day | -0.14 | 0.003 | -0.25 | 0.02 | -0.27 | 0.01 |

We then examined how redefining the severe to extremely severe stress group affected predictive performance in our machine learning models. This improved the models’ performance across all models used and performance metrics. Importantly, redefining the severe to extremely severe stress group improved both the precision and recall, suggesting that our models were indeed able to differentiate between users with no stress and those who were severely to extremely severely stressed (Table 6).

**Table 6.** Machine learning algorithm performance metrics for predicting users with no stress vs. severe to extremely severe stress.

| Machine Learning Algorithm | Precision | Recall | F1 | AUC |
| --- | --- | --- | --- | --- |
| Classification and regression trees (CART) | 0.5 | 0.67 | 0.55 | 0.55 |
| Random Forest | 0.67 | 0.67 | 0.67 | 0.62 |
| Shrinkage discriminant analysis (SDA) | 0.67 | 0.67 | 0.67 | 0.74 |
| General linear model | 0.75 | 0.69 | 0.72 | 0.68 |
| K nearest neighbour | 0.67 | 0.62 | 0.64 | 0.64 |
| Neural network | 0.93 | 0.87 | 0.83 | 0.90 |

To further visualize the differences that were predictive of stress status, we built multigraphs of passive sensing features for representative single users from both the no stress and severe to extremely severe stress groups. For the user with no stress, they visited only seven places an average time of once per day across the duration of the study. They also had a 75^th^ quantile path weight of 8.25 (Fig. 4). The severe to extremely severe user, however, visited 25 places an average of three times per day throughout the study and a 75^th^ quantile path weight of 4 (Fig. 4).

**Figure 4.**
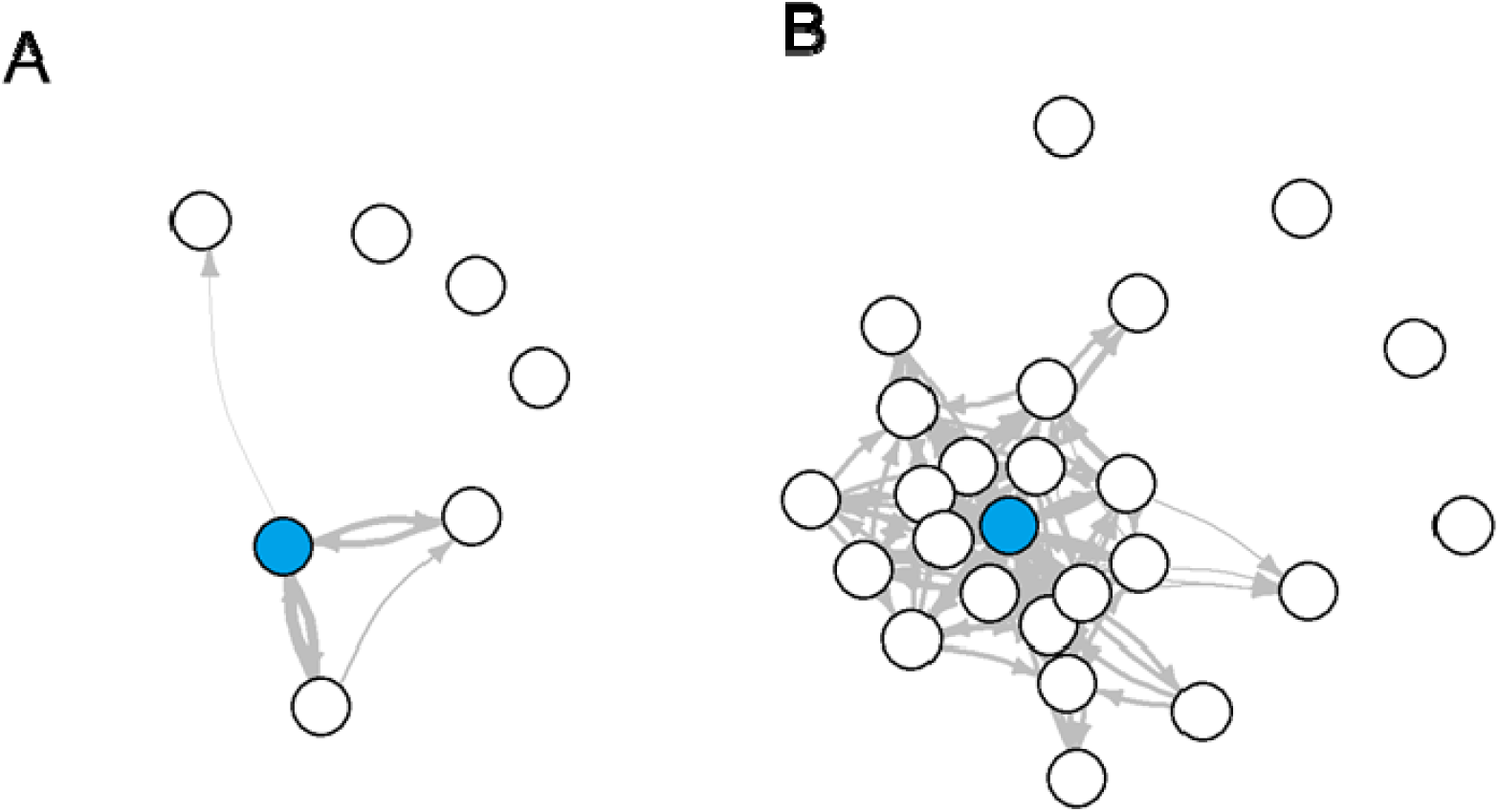
Multigraphs of representative user from the (A) no stress and (B) severe to extremely severe stress groups.

## DISCUSSION

Using a novel methodological pipeline, we showed that key features from passive sensing data served as a predictor of severe to extremely severe stress across several supervised machine learning models. Key features included number of unique nodes (locations), number of nodes visited per day (75^th^ quantile), and average number of steps per day. These passive sensing features alone were able to differentiate someone who was not stressed versus someone who was severely to extremely severely stressed.

Although two previous studies demonstrated that there was a correlation between GPS features and stress levels in university students, they focused on other features including longer distance between locations (Muller et al., 2020), evenly distributed time spent at different locations (Muller et al., 2020), and total distance travelled daily (Ben-Zeev et al., 2015). To our knowledge, this is the first paper that shows that a high number of locations, number of locations visited per day, number of steps could predict university students who were severely to extremely severely stressed. Further, this is the first study that has used supervised machine learning to demonstrate that these features can indeed predict the level of stress in university students. This is an important finding in the context of both diagnosis and treatment. First, it suggests that apps that collect passive sensing data may be used to diagnose or predict the level of stress someone is experiencing, allowing us to move away from cumbersome, and at times biased, self-report questionnaires to assess stress (Razavi, 2001). Second, our finding suggests that we can use passive sensing data to determine a mental health intervention that may be best suited to a particular user. Personalized mental health interventions have gained popularity with the recent advent of just-in-time adaptive intervention (JITAI) apps. These apps are designed to tailor interventions to the particular needs of the user based on their response to screening questionnaires (e.g. psychological self-reports) (Nahum-Shani et al., 2015). Importantly, passive sensing data analyses do not require any additional efforts on the part of the user, highlighting that apps can rapidly tailor or adjust interventions on both immediate and ongoing bases. Future research would benefit from examining whether the inclusion of the passive sensing data features can help to better tailor mental health intervention programs to the individual user.

As discussed, although there are several examples of previous work examining the relationship between GPS data and stress, these works have been limited to correlational analyses and have not fully described the data-specific methodologies. Critically, our study is the first to describe a clear methodological pipeline for extracting features from real-world passive sensing data and, using supervised machine learning, confirm that they can be used for digital phenotyping. With the growing accessibility of passive sensing data sourced from healthcare smartphone apps, there is an urgent need to establish clear methodologies that can be used and replicated by other research groups. First, this marks an important step away from black box style analyses, toward those that are both robust and reproducible. Second, clear methodological pipelines for rapid digital phenotyping from passive sensing data are essential for the success of personalized healthcare apps like JITAIs. Given that we’ve established that digital phenotyping severe to extremely severe stress is possible, there are two next steps for future research. First, our approach should be validated in a cohort of patients with severe stress-related clinical diagnoses such as major depression to establish its clinical translatability and validity. Second, future research should incorporate our methodological pipeline into a JITAI app to establish if it can improve its potential for targeted, personalized interventions.

While this is a promising first step toward using passive sensing data for digital phenotyping, there are some limitations. First, our passive sensing data was unable to digitally phenotype users with mild to moderate stress. Although we presented evidence of why this may be the case and highlighted the potential for a high level of heterogeneity in this stress group, future research should establish if there are ways to differentiate between users who are benefiting from their stress levels from those who are not. This could be done, for example, by the inclusion of additional questionnaires to assess users’ subjective experience of their current performance on tasks and ability to handle stress. From an early intervention approach, it is essential to elucidate whether digital phenotyping may be able to predict those at risk of transitioning from mild to moderate stress to more severe stress. This remains an important line of enquiry for future research. Another consideration of the present work is that the users of the Vibe Up app were more likely to be female than male, resulting in a 3:1 ratio of females to males. Although this is in line with previous research of university student mental health app users (Yang et al., 2018), it suggests that future work needs to establish whether this digital phenotyping extends to male students as well.

## Funding

This work was supported by Commonwealth of Australia Medical Research Future Fund grant MRFAI000028 Optimising treatments in mental health using AI.

### Ethics Approval

This study was approved by the University of New South Wales Human Research Ethics Committee, approval no. HC200466.

### Consent to Participate

As per the ethics approval, all participants consented to participate in this study and consented to the study team collecting data about their location and activity using sensors in their smartphone to enable research about the relationship between behaviour and distress. Participants consented for this to happen automatically for 4 weeks or until they uninstalled the study app, whichever is sooner. They also consented for their anonymized data to be stored in a databank for future research purposes. Participants could withdraw consent at any time.

### Consent for Publication

Not applicable.

### Clinical Trial Registration

This clinical trial was registered with Australia New Zealand Clinical Trials Registry (ANZCTR) on September 13^th^, 2021 under registration number ACTRN12621001223820.

### Availability of Data and Materials

Data may be made available on request and subject to the relevant governance procedures.

### Code Availability

Code may be made available on request.

### Authors Contributions

A.S. Conceived and designed the study, performed the machine learning and data analyses, and wrote the manuscript.

J.F.K., L.H., K.M., S.G., and S.V. assisted with data collection and co-wrote the manuscript.

A.W., M.S., A.S., W-Y.Z., J.H., H.C., and J.N. assisted with conceptualization and co-wrote the manuscript.

## Data Availability

Data may be made available on request and subject to the relevant governance procedures.

## REFERENCES

1. Ahuvia, I. L., Sung, J. Y., Dobias, M. L., Nelson, B. D., Richmond, L. L., London, B., & Schleider, J. L. (2022). College student interest in teletherapy and self-guided mental health supports during the COVID-19 pandemic. Journal of American College Health, 15, 1–7.

2. Andrews, G., & Slade, T. (2007). Interpreting scores on the Kessler Psychological Distress Scale (K10). Australian and New Zealand Journal of Public Health, 25(6), 494–497.

3. Bautista, J., & Schueller, S. M. (2023). Understanding the adoption and use of digital mental health apps among college students: Secondary analyses of a national survey. JMIR Mental Health., 10, e43942.

4. Beiwinkel, T., Kindermann, S., Maier, A., Kerl, C., Moock, J., Barbian, G., & Rossler, W. (2016). Using smartphones to monitor bipolar disorder symptoms: a pilot study. JMIR Mental Health., 3(1).

5. Ben-Zeev, D., Scherer, E. A., Wang, R., Xie, H., & Campbell, A. T. (2015). Next-generation psychiatric assessment: Using smartphone sensors to monitor behavior and mental health. Psychiatric Rehabilitation Journal, 38(3), 218–226.

6. Ben-Zeev, D., Wang, R., Abdullah, S., Brian, R., Scherer, E. A., Mistler, L. A., Hauser, M., Kane, J. M., Campbell, A., & Choudhury, T. (2016). Mobile behavioral sensing for outpatients and inpatients with schizophrenia. Psychiatric Services, 67(5), 558–561.

7. Beste, C., Yildiz, A., Meissner, T. W., & Wolf, O. T. (2013). Stress improves task processing efficiency in dual-tasks. Behavioral Brain Research, 252, 260–265.

8. Bianchi, R., Truchot, D., Laurent, E., Brisson, R., & Schonfeld, I. S. (2014). Is burnout solely job-related? A critical comment. Scandanavian Journal of Psychology, 55(4), 357–361.

9. Cohen, S., Janicki-Deverts, D., & Miller, G. E. (2007). JAMA, 298(14), 1685–1687.

10. Degroote, C., Schwaninger, A., Heimgartner, N., Hedinger, P., Ehlert, U., & Wirtz, P. H. (2020). Acute stress improves concentration performance. Experimental Psychology 67(2).

11. Galambos, C., Skubic, M., Wang, S., & Rantz, M. (2013). Management of dementia and depression utilizing in-home passive sensor data. Gerontechnology, 11(3), 457–468.

12. Hamaideh, S. H. (2011). Stressors and reactions to stressors among university students. International Journal of Social Psychiatry, 57(1), 69–80.

13. Holtz, B., McCarroll, A., & Mitchell, K. (2020). Perceptions and Attitudes Toward a Mobile Phone App for Mental Health for College Students: Qualitative Focus Group Study. JMIR Formative Research, 4(8).

14. Huckvale, K., Hoon, L., Stech, E., Newby, J., Zheng, W.-Y., Han, J., Vasa, R., Gupta, S., Barnett, S., Senadeera, M., Cameron, S., Kurniawan, S., Agarwal, A., Kupper, J. F., Asbury, J., Willie, D., Grant, A., Cutler, H., Parkinson, B., Christensen, H. (2023). Protocol for a bandit-based response adaptive trial to evaluate the effectiveness of brief self-guided digital interventions for reducing psychological distress in university students: the Vibe Up study BMJ Open, 13(4), e066249.

15. Kern, A., Hong, V., Song, J., Lipson, S. K., & Eisenberg, D. (2018). Mental health apps in a college setting: openness, usage, and attitudes. mHealth, 4:20.

16. Lovibond, S. H., & Lovibond, P. F. (1995). Manual for the depression, anxiety and stress scales (DASS). Psychology Foundation.

17. Lu, L. (1994). University transition: Major and minor life stressors, personality characteristics and mental health. Psychological Medicine, 24(1), 81–87.

18. Melamed, S., Kushnir, T., & Shirom, A. (1992). Burnout and risk factors for cardiovascular diseases Behavioral Medicine, 18(2), 53–60.

19. Melcher, J., Camacho, E., Lagan, S., & Torous, J. (2022). College student engagement with mental health apps: analysis of barriers to sustained use. Journal of American College Health, 70:6, 1819–1825.

20. Montagni, I., Cariou, T., Feuillet, T., Langlois, E., & Tzourio, C. (2018). Exploring digital health use and opinions of university students: Field survey study. JMIR Mhealth Uhealth, 6(3), e65.

21. Muller, S. R., Peters, H., Matz, S. C., Wang, W., & Harari, G. M. (2020). Investigating the relationship between mobility behaviours and indicators of subjective well-being using smartphone-based experience sampling and GPS tracking. European Journal of Personality 34, 714–732.

22. Muscatell, K. A., Slavich, G. M., Monrow, S. M., & Gotlib, I. H. (2009). Stressful life events, chronic difficulties, and the symptoms of clinical depression. Journal of Nervous Mental Disorders, 197(3), 154–160.

23. Nahum-Shani, I., Hekler, E. B., & Spruijt-Metz, D. (2015). Building health behavior models to guide the development of just-in-time adaptive interventions: A pragmatic framework. Health Psychology, 34(Supp), 1209–1219.

24. Neary, M., & Schueller, S. M. (2018). State of the field of mental health apps. Cognitive and Behavioral Practice, 25(4), 531–537.

25. Parkitny, L., & McAuley, J. (2010). The depression anxiety stress scale (DASS). Journal of Physiotherapy, 56, 204.

26. Razavi, T. (2001). Self-report measures: An overview of concerns and limitations of questionnaire use in occupational stress research.

27. Ribeiro, Í. J.S., Pereira, R., Freire, I. V., de Oliveira, B. G., Casotti, C. A., & Boery, E. N. (2018). Stress and Quality of Life Among University Students: A Systematic Literature Review. Health Professions Education, 4(2), 70–77.

28. Saeb, S., Zhang, M., Karr, C. J., Schueller, S. M., Corden, M. E., Kording, K. P., & Mohr, D. C. (2015). Mobile phone sensor correlates of depressive symptom severity in daily-life behavior: an exploratory study. Journal of Medical Internet Research, 17(7).

29. Sawatzky, R. G., Ratner, P. A., Richardson, C. G., Washburn, C., Sudmant, W., & Mirwaldt, P. (2012). Stress and depression in students the mediating role of stress management self-efficacy. Nursing Research, 61(1), 13–21.

30. Sharp, J., & Theiler, S. (2018). A Review of Psychological Distress Among University Students: Pervasiveness, Implications and Potential Points of Intervention. International Journal for the Advancement of Counselling, 40, 193–212.

31. Steptoe, A., & Kivimäki, M. (2013). Stress and cardiovascular disease: an update on current knowledge. Annual Review of Public Health, 34, 337–354.

32. van Spijker, B. A. J., Batterham, P. J., Calear, A. L., Farrer, L., Christensen, H., Reynolds, J., & Kerkhof, J. F. M. (2014). The suicidal ideation attributes scale (SIDAS): Community-based validation study of a new scale for the measurement of suicidal ideation. Suicide and Life-Threatening Behavior, 44(4), 408–419.

33. Verger, P., Combes, J.-B., Kovess-Masfety, V., Choquet, M., Guagliardo, V., Rouillon, F., & Peretti-Wattel, P. (2008). Psychological distress in first year university students: Socioeconomic and academic stressors, mastery, and social support in young men and women. Social Psychiatry and Psychiatric Epidemiology, 44, 643–650.

34. Weber, A., & Jackel-Reinhard, A. (2000). Burnout syndrome: A disease of modern societies? Occupational Medicine, 50(7), 512–517.

35. Winsa, B., Adami, H. O., Bergström, R., Gamstedt, A., Dahlberg, P. A., Adamson, U., Jansson, R., & Karlsson, A. (1991). Stressful life events and Graves’ disease. Lancet, 338, 1475–1479.

36. Yang, S.-Y., Lin, C.-Y., Huang, Y.-C., & Chang, J.-H. (2018). Gender differences in the association of smartphone use with the vitality and mental health of adolescent students. Journal of American College Health, 66(7).

37. Yuen, E. Y., Liu, W., Karatsoreos, I., Feng, J., McEwen, B. S., & Yan, Z. (2009). Acute stress enhances glutamatergic transmission in prefrontal cortex and facilitates working memory. PNAS, 106 (33), 14075–14079.

